# Protocol for a mixed methods process evaluation of the Smoking Treatment Optimisation in Pharmacies (STOP) trial

**DOI:** 10.1101/2020.03.17.20037499

**Authors:** Sandra Jumbe, Vichithranie Madurasinghe, Colin Houlihan, Samantha L Jumbe, Wai-Yee James, Stephanie JC Taylor, Robert Walton

## Abstract

**Introduction:** Assessing the fidelity of complex behavioural interventions and examining the contextual reasons why such interventions succeed, or fail are important activities but challenging and rarely reported. The Smoking Treatment Optimisation in Pharmacies (STOP) trial is a cluster randomised trial evaluating the effectiveness of a complex intervention to optimise the National Health Service (NHS) Stop Smoking Service delivered in community pharmacies. This complex intervention comprises a training package for pharmacy staff involving motivational interviewing and communication skills aimed at increasing smoking cessation knowledge and proactive client engagement. We report on a process evaluation which was planned alongside the trial to offer findings that will assist in the interpretation of the main trial results and help inform potential implementation in community pharmacy settings on a wider scale.

**Methods and analysis:** Quantitative data on recruitment and retention process of pharmacies, pharmacy staff and service users has been collected during the trial along with data on dose and fidelity of the intervention delivery from participating intervention arm pharmacies to identify potential implementation issues. Simulated client data on behaviour change skills and display of intervention materials from both control and intervention pharmacies is being assessed. These data will be combined with qualitative data; including adviser-smoker consultation recordings that provide a snapshot of behaviour skills delivery by stop smoking advisers and semi-structured interviews with pharmacy staff and services users from the intervention arm.

**Discussion:** Published protocols for process evaluations of complex health interventions are still rare despite increasing funding for this work to facilitate understanding of trial outcomes from an implementation perspective. This mixed methods protocol will contribute to the developing literature around the conduct of process evaluation and the value they add to health services research.

***Trial registration number*** ISRCTN16351033.

**Strengths and limitations of this study:**

- A planned mixed methods process evaluation that draws together data from different sources to help explain the trial results and establish the feasibility of scaling this complex intervention up in community pharmacy settings.
- A strength is the use of a previously tested mystery shopping method to assess fidelity of skills performance at the pharmacy counter
- The process evaluation relies on willing pharmacy staff and service users involved in the trial to collect some of the data, which may introduce bias.
- This paper also provides a detailed example of how to use the MRC framework for process evaluation of complex interventions to design an extensive process evaluation within trial settings.

## Introduction

The Smoking Treatment Optimisation in Pharmacies (STOP) trial is a cluster randomised trial to evaluate the effectiveness of a complex intervention to optimise the NHS Stop Smoking Service delivered in community pharmacies [1]. The trial involves 60 pharmacies across London, Coventry and Cwm Taf in Wales. Thirty of these pharmacies are randomised to the intervention group, where consenting pharmacy staff are invited to a half-day session which includes motivational interviewing and communication skills training to improve smoker engagement and equip them as facilitators for health behaviour change [2, 3]. Intervention pharmacies are also given badges, a poster and a desk calendar to use in their pharmacy environment as prompts for more proactive smoking cessation dialogue with potential clients. The control pharmacies continue to deliver their service as normal over the trial duration. The primary outcome is rate of smoker recruitment into the NHS stop smoking service [1]. The trial, along with this process evaluation, was funded the NIHR Programme Grant for Applied Research [RP-PG-0609-10181]. In this paper we describe the protocol for a process evaluation which aims to evaluate how the intervention is implemented, the mechanisms by which it works and how these may be affected by the context in which the intervention is operating [4].

Process evaluations can be used to assess the fidelity and quality of implementation of an intervention, clarify causal mechanisms and identify contextual factors associated with variation in outcomes [4]. They are recommended to contextualise the results of randomised controlled trials to answer key questions about why an intervention has failed or succeeded and how it was implemented [5]. Process evaluations are particularly relevant to complex, multi-faceted interventions such as STOP that involve multiple targets (for example, pharmacy staff and patients/ service users), have various active components and where their complexity makes it difficult to measure their effects [2, 6].

In our previous STOP qualitative and pilot studies, stop smoking advisers felt they lacked the interpersonal skills necessary to engage well with smokers and help them to quit [7]. Advisers suggested regular skills training for all pharmacy staff, including staff not formally trained as stop smoking advisers, to improve uptake to their stop smoking service [2-3, 7-8]. Despite expanding the intervention training to include staff members that were not directly involved in delivering the pharmacy smoking cessation service in the main trial, we noticed a high level of staff turnover in a majority of participating trial pharmacies which may affect the reach/implementation of the intervention. Specifically, even if high levels of attendance to the intervention training are achieved, the effectiveness of the behavioural aspect of the intervention may be constrained/ diluted over time if many intervention trained staff leave the pharmacy for whatever reason.

Variations in organisational structures of pharmacies and commissioning systems also caused complications in the process of the trial. We noted that the routes to receiving payment (e.g. smoker registration vs 4 week quit outcome) and payments offered (total amount received per smoker plus bonus for specific groups) for pharmacies providing the NHS Stop Smoking Service differed in London, Coventry and Wales depending on each service commissioner’s policy [9]. These differences also extend into data collection where the type of routine data that is collected in each geographical location varied because of the different computer systems used. London and Coventry for example both used electronic portals to track and upload claims and patient data, but did not collect the exact same data fields. On the other extreme Wales relied more heavily on paper records which the STOP team used to collect and manually transcribe onto an electronic spreadsheet.

The above challenges highlighted a need to conduct a pragmatic process evaluation that allows us to understand better the contextual factors and causal mechanisms in community pharmacy settings that could affect implementation of the STOP intervention during the trial and subsequent trial outcomes. The process evaluation will support quantitative data collected from the 60 participating pharmacies across three regions – London, Coventry and Cwm Taf. The evaluation will enable us to place the pharmacies in context, assess fidelity of the intervention, quantify the intervention dose delivered, the intervention dose received and the extent to which the target population participated in the intervention (including recruitment rates for pharmacies, staff and smokers).

The design of the process evaluation is underpinned by the Medical Research Council guidance [4] and the theoretical framework used to develop the STOP intervention [2, 3]. Specifically, development of this protocol is based on the key components of process evaluation proposed by Moore and colleagues [4] which are: context, reach, dose delivered, dose received, fidelity, implementation and recruitment [10]. From a theoretical perspective, outputs from the activities outlined in the final STOP logic model (Figure 1) alongside the STOP programme theories will be used to identify the causal processes through which change comes about as a result of a programme’s strategies and action [11]. In addition, we decided that it was important to include the views and opinions of participants, including pharmacy staff who attended the STOP intervention training and consenting STOP Trial service users, who used the stop smoking service at participating pharmacies. Their experiences, their attitudes to the intervention and how they think the intervention could be improved will play an important part in interpreting the outcome data and developing subsequent implementation. This process evaluation protocol establishes how we will evaluate the fidelity of implementation of the STOP intervention, clarify causal mechanisms and identify factors associated with variation in the STOP Trial outcomes [1].

**Figure 1:**
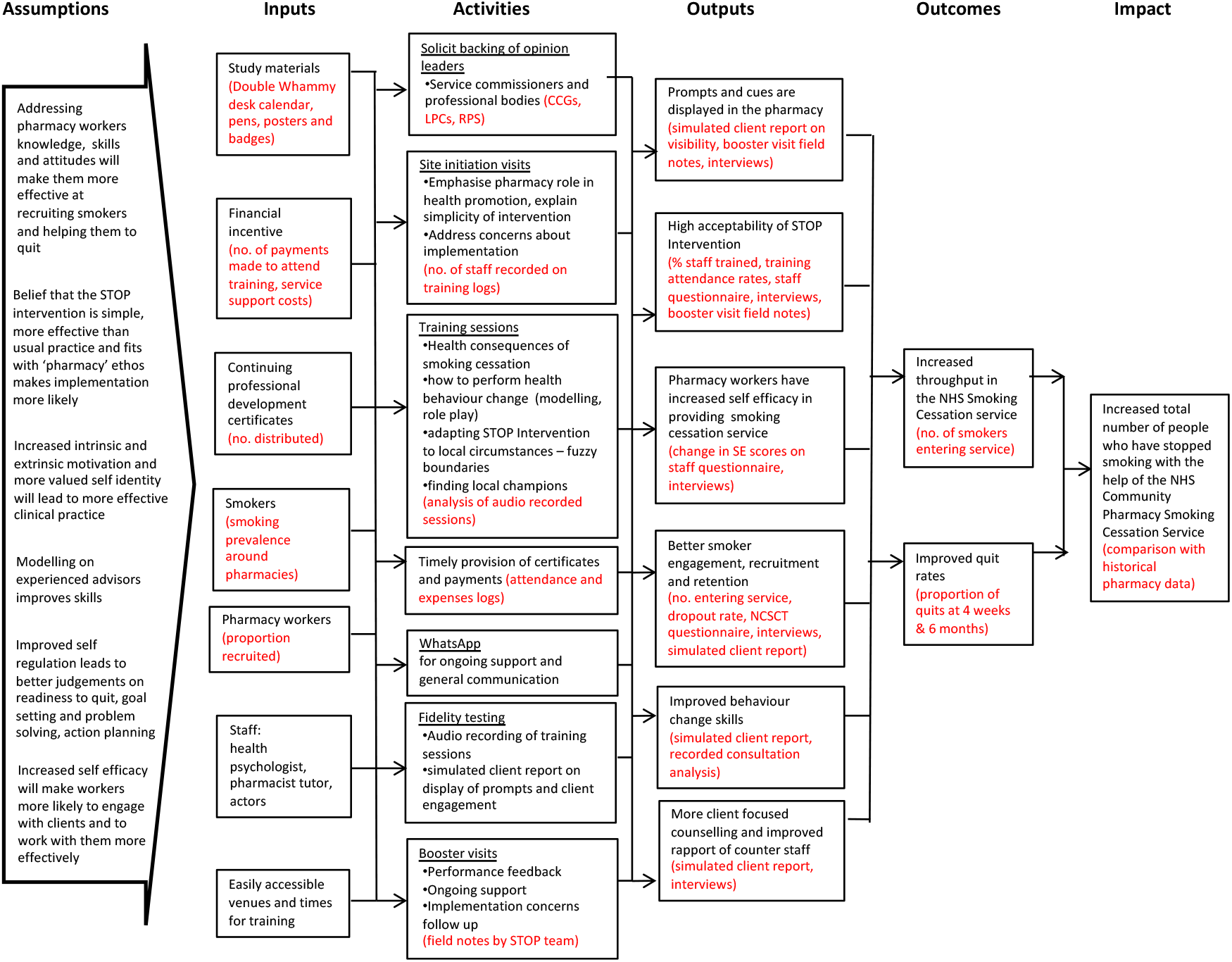
STOP Trial Logic model

The specific process evaluation aims for the STOP Trial are;

1. To provide findings that will assist in the interpretation of the clinical trial results, particularly understanding how and in what circumstances the STOP Intervention improves smoker uptake (or not)
2. To inform potential implementation of the STOP training intervention in community pharmacy settings on a wider scale

## Methods

### Design

This process evaluation will use quantitative and qualitative data. The use of mixed methods research is increasingly common in health services research and evaluation [12]. Specific to process evaluations, a mixed method approach can help researchers to explore apparent discrepancies between findings [12-13]. There are practicalities to this methodological approach that require careful forward planning including how each method will complement the other, and which data collection methods to use to achieve the project’s goals [13]. Quantitative data (e.g. intervention dosage, pharmacy staff client engagement scores from simulated clients; facilitator training adherence scores) will be complemented by qualitative data from pharmacy staff and service user interviews, intervention training attendees’ feedback, plus field notes from pharmacy booster visits and simulated client visits to illuminate why the intervention was effective or not [3, 13].

### Process evaluation objectives

➢ Collect quantitative data on number of pharmacies recruited, and pharmacy staff consented to the STOP Trial, records of pharmacy staff trained in informed consent in research and trial related processes during site initiation visits. Include reported numbers of staff employed by participating pharmacies to assess reach
➢ Collect quantitative data on the proportion of pharmacy staff invited and attending training with reasons for non-attendance from the intervention arm. Data on training venues will also be collected
➢ Collate and analyse quantitative data from simulated clients regarding display of intervention materials and client engagement by pharmacy staff at the counter. This data will also include field notes to be analysed qualitatively
➢ Analyse audio recordings of STOP training sessions to assess fidelity of intervention delivery by training facilitators
➢ Assess acceptability of the STOP intervention by interviewing pharmacy staff about their views of the STOP training, and perceived impact of the intervention on their NHS stop smoking service. These interviews will be transcribed, then later analysed qualitatively alongside pharmacy staff post training feedback data collected after each training session and 5 months later.
➢ Collate data on service user satisfaction of the NHS SSS from the NCSCT questionnaire
➢ Carry out interviews with consenting service users on their views of the stop smoking service from participating pharmacies
➢ Collate data on number of booster visits conducted in intervention pharmacies, and analyse field notes, describing contextual factors reported by pharmacy staff that hindered or facilitated implementation of the STOP intervention in their environment
➢ Interpret key outputs and results to provide informative descriptive reports and statistical analyses
➢ Disseminate comprehensive process evaluation outcomes report through peer reviewed articles, and presentations at academic conferences and stakeholder events

Figure 2 provides a summary of key process evaluation components in accordance with MRC guidance, associated theoretical assumptions (see Figure 1), and how these will be measured within data collected.

**Figure 2:**
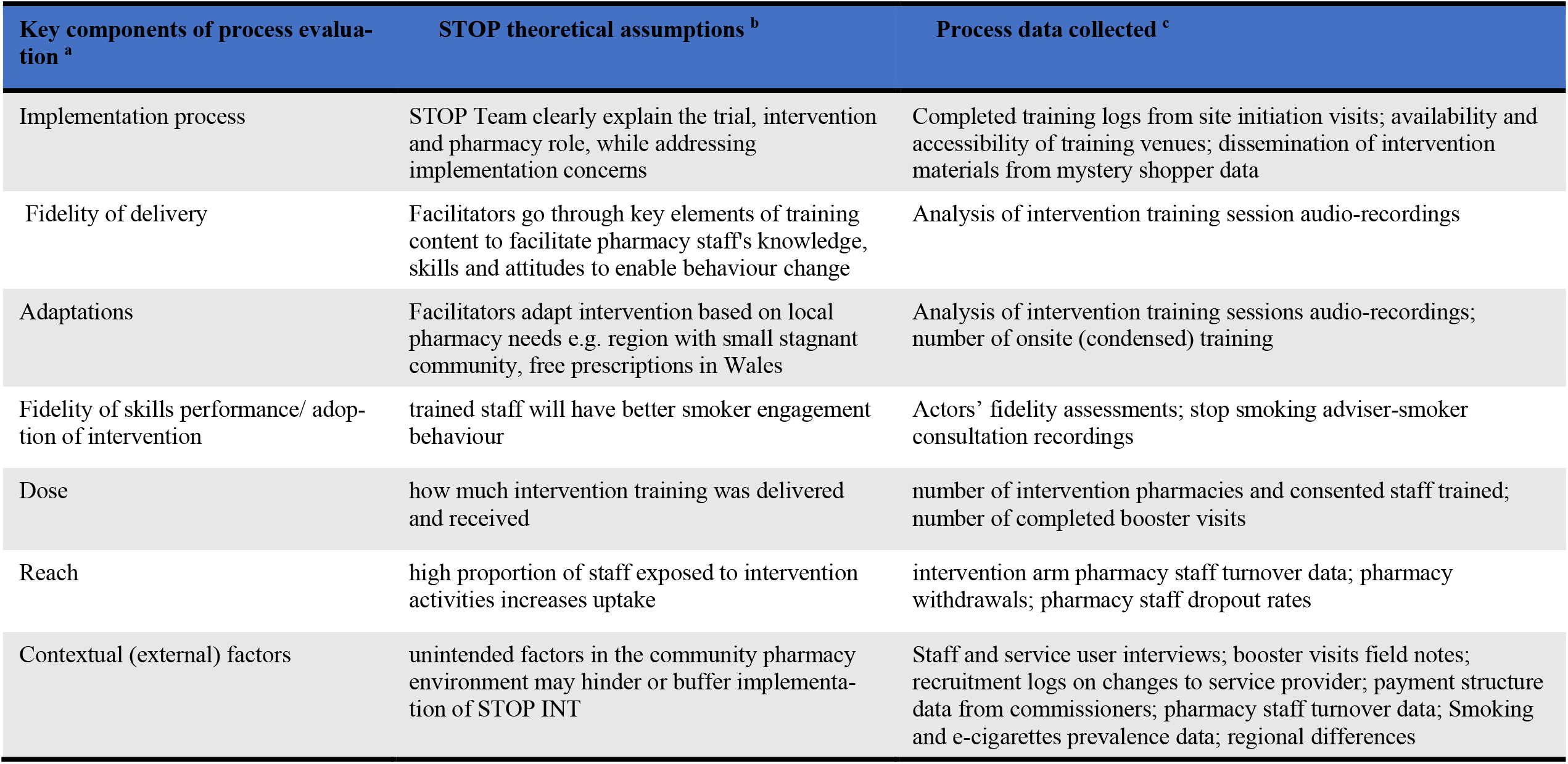
Key MRC process evaluation components, associated STOP process evaluation theoretical assumptions and data ^a^ from MRC guidance (Moore et al, 2015) ^b^ based on STOP Logic model ^c^ outlined in the STOP Trial protocol (Sohanpal et al, 2019)

### Data Collection Methods

Table 1 outlines the methods used to collect specific STOP Trial data for this process evaluation.

**Table 1:**
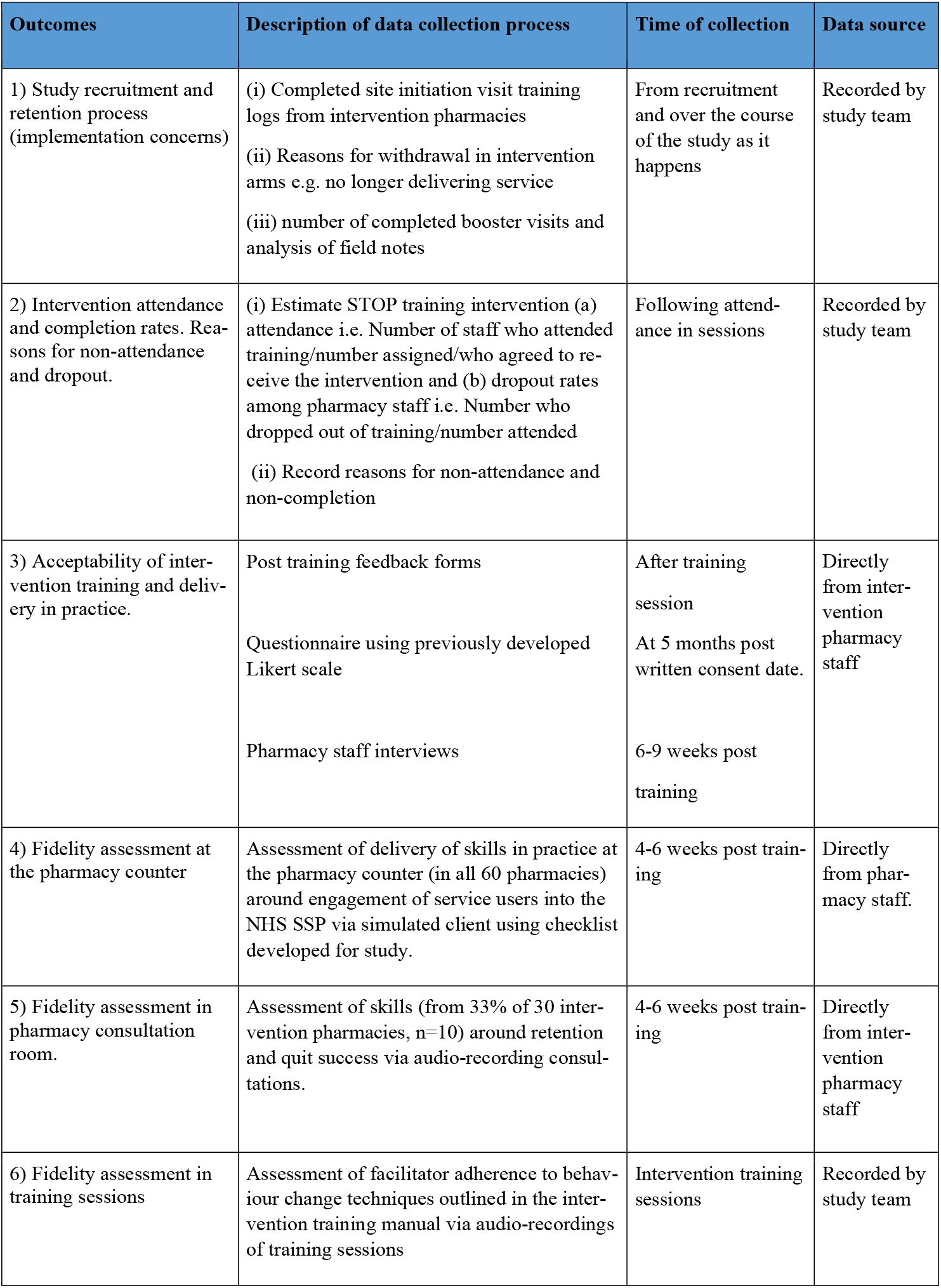
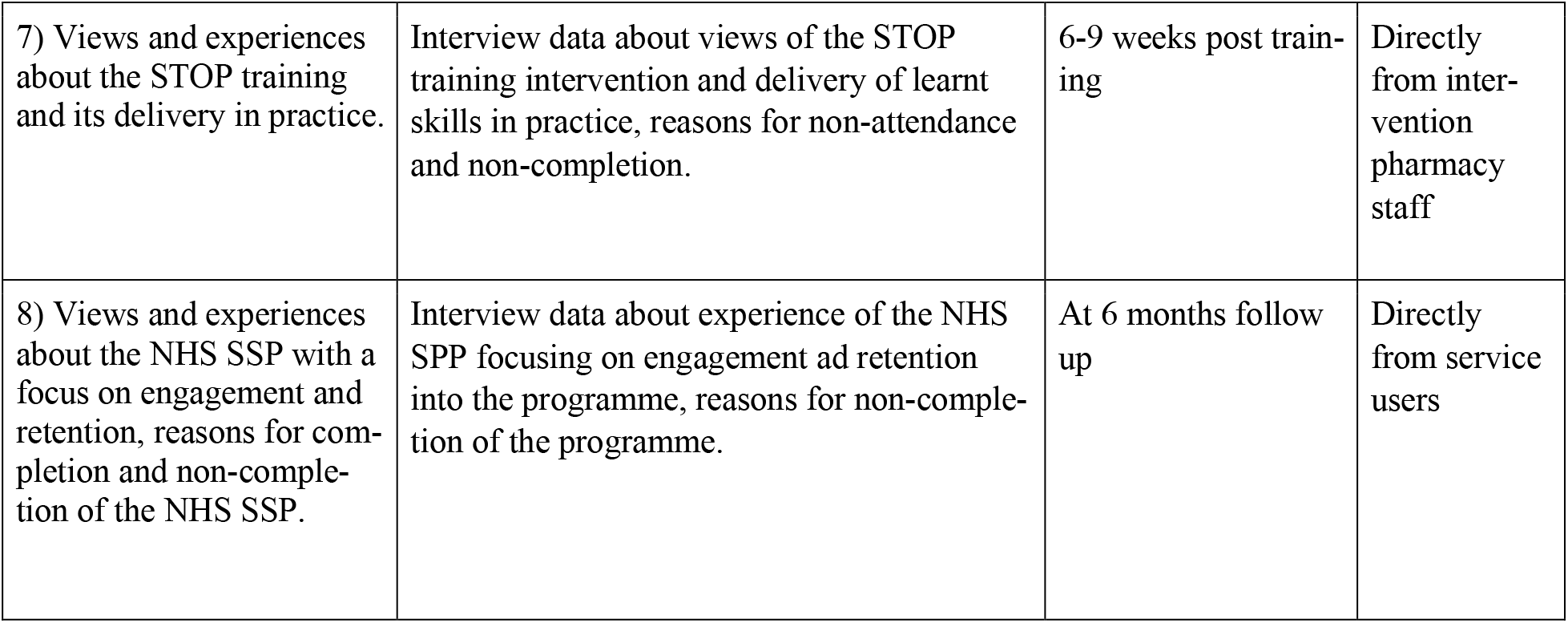
Summary of STOP Trial process data collection

| Outcomes | Description of data collection process | Time of collection | Data source |
| --- | --- | --- | --- |
| 1) Study recruitment and retention process (implementation concerns) | (i) Completed site initiation visit training logs from intervention pharmacies<br>(ii) Reasons for withdrawal in intervention arms e.g. no longer delivering service<br>(iii) number of completed booster visits and analysis of field notes | From recruitment and over the course of the study as it happens | Recorded by study team |
| 2) Intervention attendance and completion rates. Reasons for non-attendance and dropout. | (i) Estimate STOP training intervention (a) attendance i.e. Number of staff who attended training/number assigned/who agreed to receive the intervention and (b) dropout rates among pharmacy staff i.e. Number who dropped out of training/number attended<br>(ii) Record reasons for non-attendance and non-completion | Following attendance in sessions | Recorded by study team |
| 3) Acceptability of intervention training and delivery in practice. | Post training feedback forms<br>Questionnaire using previously developed Likert scale<br>Pharmacy staff interviews | After training session<br>At 5 months post written consent date.<br>6-9 weeks post training | Directly from intervention pharmacy staff |
| 4) Fidelity assessment at the pharmacy counter | Assessment of delivery of skills in practice at the pharmacy counter (in all 60 pharmacies) around engagement of service users into the NHS SSP via simulated client using checklist developed for study. | 4-6 weeks post training | Directly from pharmacy staff. |
| 5) Fidelity assessment in pharmacy consultation room. | Assessment of skills (from 33% of 30 intervention pharmacies, n=10) around retention and quit success via audio-recording consultations. | 4-6 weeks post training | Directly from intervention pharmacy staff |
| 6) Fidelity assessment in training sessions | Assessment of facilitator adherence to behaviour change techniques outlined in the intervention training manual via audio-recordings of training sessions | Intervention training sessions | Recorded by study team |
| 7) Views and experiences about the STOP training and its delivery in practice. | Interview data about views of the STOP training intervention and delivery of learnt skills in practice, reasons for non-attendance and non-completion. | 6-9 weeks post training | Directly from intervention pharmacy staff |
| 8) Views and experiences about the NHS SSP with a focus on engagement and retention, reasons for completion and non-completion of the NHS SSP. | Interview data about experience of the NHS SPP focusing on engagement and retention into the programme, reasons for non-completion of the programme. | At 6 months follow up | Directly from service users |

Fidelity assessment will be conducted in three ways:

1. Audio-recordings of STOP pharmacy staff training sessions
2. Assessment of prompts and client engagement with staff at the pharmacy counter using simulated clients
3. Audio-recordings of advisor-smoker consultations

Audio-recordings of STOP pharmacy staff training sessions

Audio-recordings of training sessions will only be obtained if the group provides verbal consent to be recorded. Specifically, before starting each training session, the training facilitators (or study team) will ask attendees being trained if they are happy for their session to be recorded. Facilitators will explain the reason for this i.e. to assess adherence to intervention content being delivered according to the STOP intervention training manual. Content in this training manual has been previously coded into component behaviour change techniques (BCTs) by two health psychologists from the study team (see table 2) with previous training and experience in coding using the BCT taxonomy [2, 3].

**Table 2:**
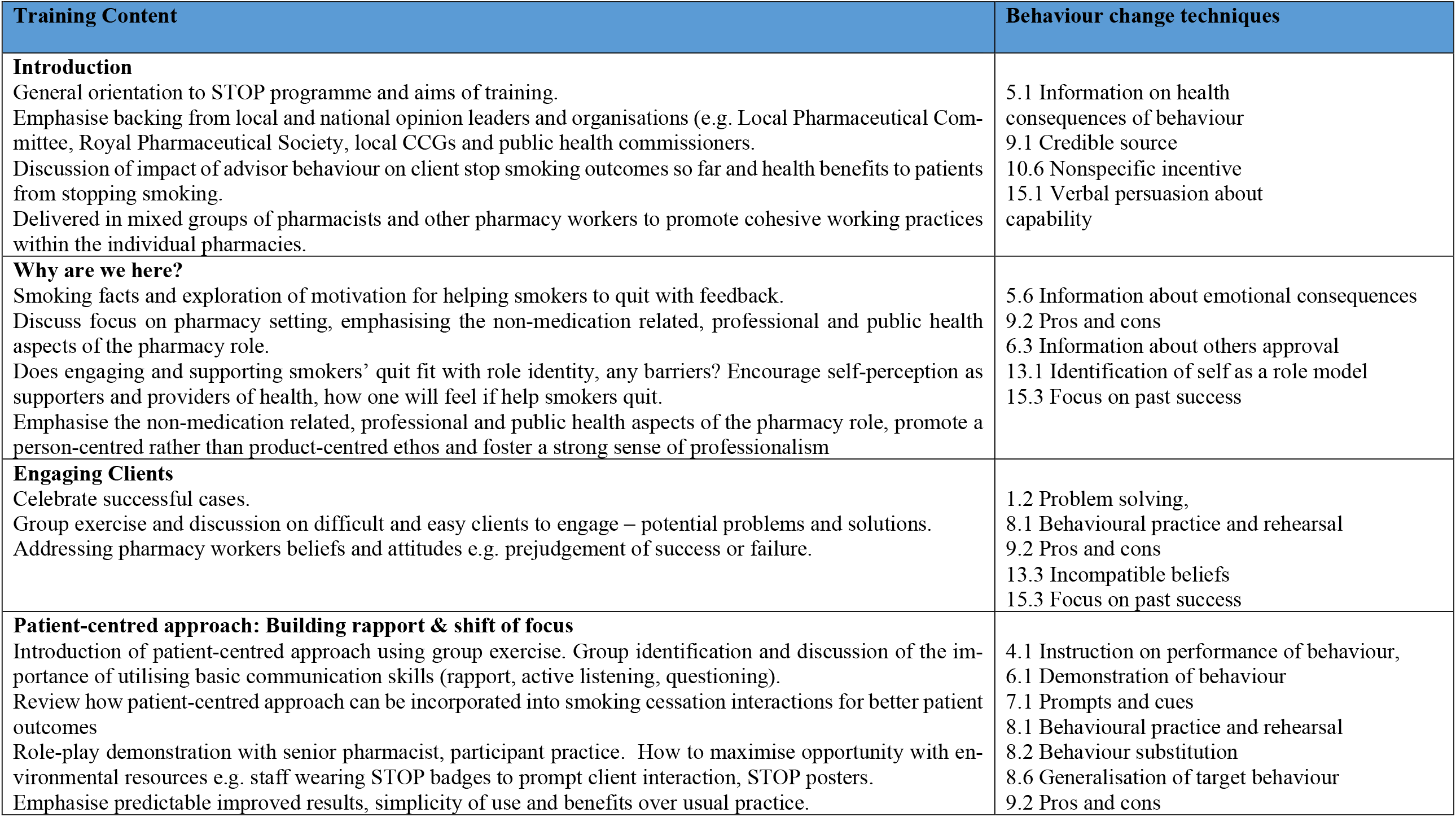
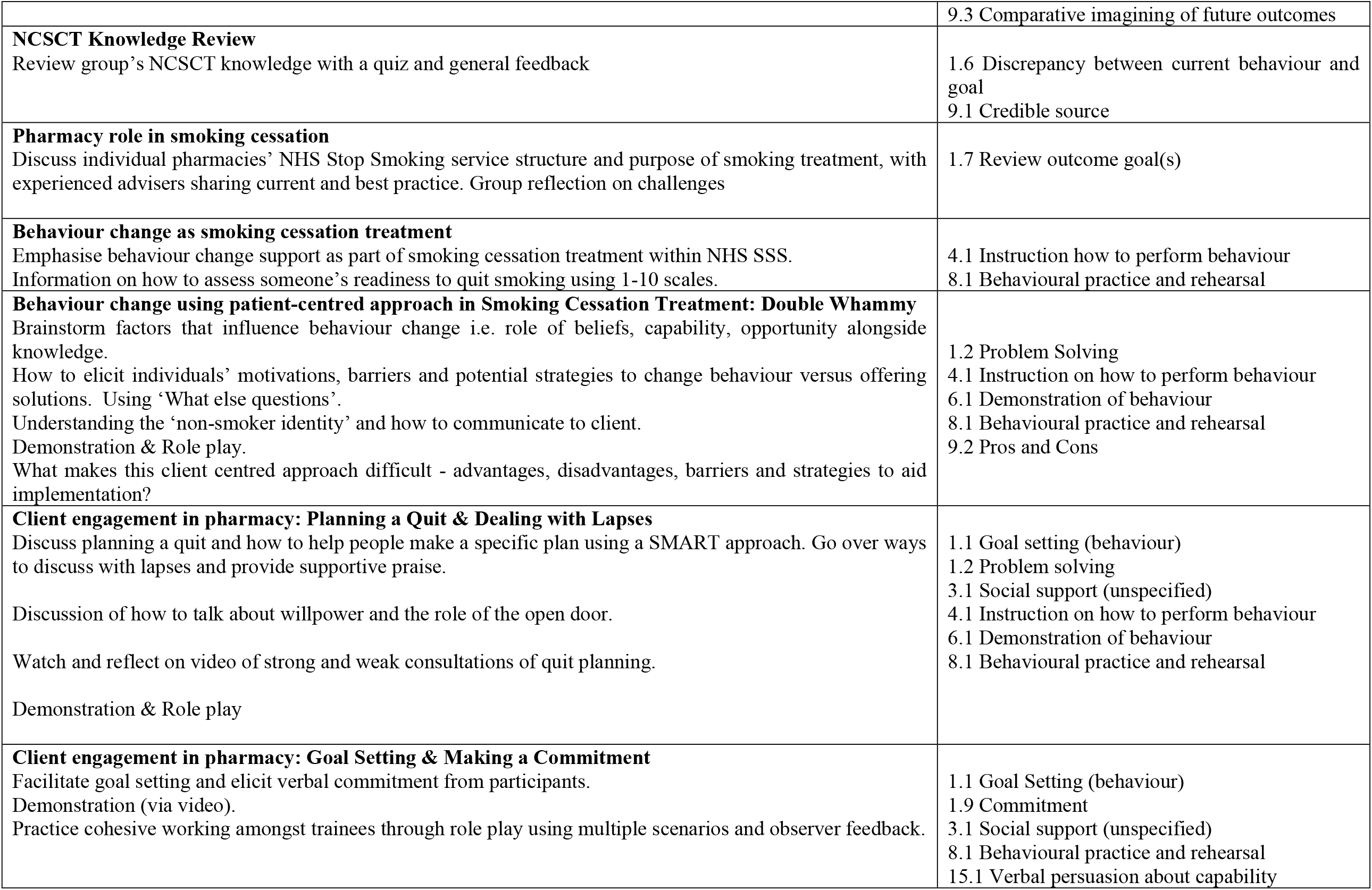
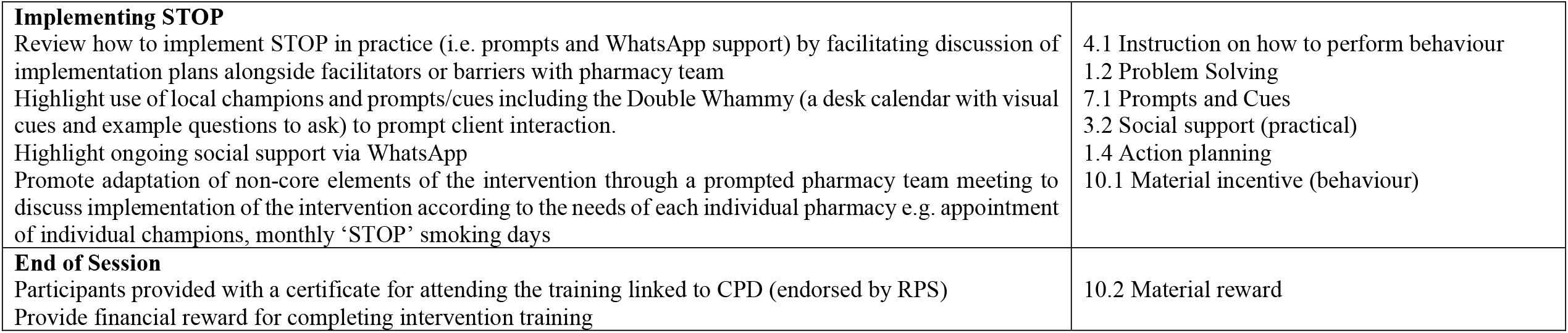
STOP Trial Intervention Training Content

| Training Content | Behaviour change techniques |
| --- | --- |
| **Introduction**<br>General orientation to STOP programme and aims of training.<br>Emphasise backing from local and national opinion leaders and organisations (e.g. Local Pharmaceutical Committee, Royal Pharmaceutical Society, local CCGs and public health commissioners.<br>Discussion of impact of advisor behaviour on client stop smoking outcomes so far and health benefits to patients from stopping smoking.<br>Delivered in mixed groups of pharmacists and other pharmacy workers to promote cohesive working practices within the individual pharmacies. | 5.1 Information on health consequences of behaviour<br>9.1 Credible source<br>10.6 Nonspecific incentive<br>15.1 Verbal persuasion about capability |
| **Why are we here?**<br>Smoking facts and exploration of motivation for helping smokers to quit with feedback.<br>Discuss focus on pharmacy setting, emphasising the non-medication related, professional and public health aspects of the pharmacy role.<br>Does engaging and supporting smokers' quit fit with role identity, any barriers? Encourage self-perception as supporters and providers of health, how one will feel if help smokers quit.<br>Emphasise the non-medication related, professional and public health aspects of the pharmacy role, promote a person-centred rather than product-centred ethos and foster a strong sense of professionalism | 5.6 Information about emotional consequences<br>9.2 Pros and cons<br>6.3 Information about others approval<br>13.1 Identification of self as a role model<br>15.3 Focus on past success |
| **Engaging Clients**<br>Celebrate successful cases.<br>Group exercise and discussion on difficult and easy clients to engage – potential problems and solutions.<br>Addressing pharmacy workers beliefs and attitudes e.g. prejudgement of success or failure. | 1.2 Problem solving,<br>8.1 Behavioural practice and rehearsal<br>9.2 Pros and cons<br>13.3 Incompatible beliefs<br>15.3 Focus on past success |
| **Patient-centred approach: Building rapport & shift of focus**<br>Introduction of patient-centred approach using group exercise. Group identification and discussion of the importance of utilising basic communication skills (rapport, active listening, questioning).<br>Review how patient-centred approach can be incorporated into smoking cessation interactions for better patient outcomes<br>Role-play demonstration with senior pharmacist, participant practice. How to maximise opportunity with environmental resources e.g. staff wearing STOP badges to prompt client interaction, STOP posters.<br>Emphasise predictable improved results, simplicity of use and benefits over usual practice. | 4.1 Instruction on performance of behaviour,<br>6.1 Demonstration of behaviour<br>7.1 Prompts and cues<br>8.1 Behavioural practice and rehearsal<br>8.2 Behaviour substitution<br>8.6 Generalisation of target behaviour<br>9.2 Pros and cons |
|  | 9.3 Comparative imagining of future outcomes |
| **NCSCT Knowledge Review**<br>Review group's NCSCT knowledge with a quiz and general feedback | 1.6 Discrepancy between current behaviour and goal<br>9.1 Credible source |
| **Pharmacy role in smoking cessation**<br>Discuss individual pharmacies' NHS Stop Smoking service structure and purpose of smoking treatment, with experienced advisers sharing current and best practice. Group reflection on challenges | 1.7 Review outcome goal(s) |
| **Behaviour change as smoking cessation treatment**<br>Emphasise behaviour change support as part of smoking cessation treatment within NHS SSS.<br>Information on how to assess someone's readiness to quit smoking using 1-10 scales. | 4.1 Instruction how to perform behaviour<br>8.1 Behavioural practice and rehearsal |
| **Behaviour change using patient-centred approach in Smoking Cessation Treatment: Double Whammy**<br>Brainstorm factors that influence behaviour change i.e. role of beliefs, capability, opportunity alongside knowledge.<br>How to elicit individuals' motivations, barriers and potential strategies to change behaviour versus offering solutions. Using 'What else questions'.<br>Understanding the 'non-smoker identity' and how to communicate to client.<br>Demonstration & Role play.<br>What makes this client centred approach difficult - advantages, disadvantages, barriers and strategies to aid implementation? | 1.2 Problem Solving<br>4.1 Instruction on how to perform behaviour<br>6.1 Demonstration of behaviour<br>8.1 Behavioural practice and rehearsal<br>9.2 Pros and Cons |
| **Client engagement in pharmacy: Planning a Quit & Dealing with Lapses**<br>Discuss planning a quit and how to help people make a specific plan using a SMART approach. Go over ways to discuss with lapses and provide supportive praise.<br>Discussion of how to talk about willpower and the role of the open door.<br>Watch and reflect on video of strong and weak consultations of quit planning.<br>Demonstration & Role play | 1.1 Goal setting (behaviour)<br>1.2 Problem solving<br>3.1 Social support (unspecified)<br>4.1 Instruction on how to perform behaviour<br>6.1 Demonstration of behaviour<br>8.1 Behavioural practice and rehearsal |
| **Client engagement in pharmacy: Goal Setting & Making a Commitment**<br>Facilitate goal setting and elicit verbal commitment from participants.<br>Demonstration (via video).<br>Practice cohesive working amongst trainees through role play using multiple scenarios and observer feedback. | 1.1 Goal Setting (behaviour)<br>1.9 Commitment<br>3.1 Social support (unspecified)<br>8.1 Behavioural practice and rehearsal<br>15.1 Verbal persuasion about capability |
| **Implementing STOP**<br>Review how to implement STOP in practice (i.e. prompts and WhatsApp support) by facilitating discussion of implementation plans alongside facilitators or barriers with pharmacy team<br>Highlight use of local champions and prompts/cues including the Double Whammy (a desk calendar with visual cues and example questions to ask) to prompt client interaction.<br>Highlight ongoing social support via WhatsApp<br>Promote adaptation of non-core elements of the intervention through a prompted pharmacy team meeting to discuss implementation of the intervention according to the needs of each individual pharmacy e.g. appointment of individual champions, monthly ‘STOP’ smoking days | 4.1 Instruction on how to perform behaviour<br>1.2 Problem Solving<br>7.1 Prompts and Cues<br>3.2 Social support (practical)<br>1.4 Action planning<br>10.1 Material incentive (behaviour) |
| **End of Session**<br>Participants provided with a certificate for attending the training linked to CPD (endorsed by RPS)<br>Provide financial reward for completing intervention training | 10.2 Material reward |

Two qualitative researchers (independent from the study team) will independently listen to each recording and rate (coded as 0 (NO) or 1 (YES) whether the facilitator applied all specified BCTs outlined in the manual,[14] and learnt during the training in practice. They will then subsequently meet to review their ratings and resolve any discrepancies along with the process evaluation team/ study health psychologist. If the researchers rate the BCTs within a section of the training the same, agreement will be registered. Where one researcher verifies a BCT and the other does not, disagreement will be registered.

### Assessing client engagement in pharmacies using simulated clients

Actors will visit each pharmacy and present at the counter with different smoking related scenarios which have been designed to present an opportunity for the pharmacy staff to engage the potential client with the smoking cessation service. Details on the actors’ training and the specific scenarios used for this process and how the actors rate client engagement are outlined in a previously published paper [3]. In brief, after completing their smoking related scenario with a member of staff at the pharmacy counter, the simulated client will rate the interaction, providing a subjective assessment of skills delivery. They will also make a note of any visible NHS stop smoking service or STOP Trial promotional materials e.g. STOP badges, desktop calendars. Each actor will complete this assessment using a checklist after each pharmacy visit which has the option of adding brief notes [3]. All actors will receive training in this mystery shopper approach by the study team. The data collected will be analysed using descriptive statistics.

### Audio-recording of adviser-smoker consultations

Over the trial duration, the study team will ask stop smoking advisers from participating pharmacies in the intervention arm to record one or two of their consultations with consenting smokers who have joined their stop smoking service. These recordings will be used to assess enactment and retention of behaviour change skills from advisers. Advisers who agree to do this will be given an encrypted audio recorder to use for this purpose. The data collected will undergo Roter interaction analysis which is a method widely used to code medical interactions worldwide [15]. The Roter interaction analysis system (RIAS) emerged in the late 90s when research studies looking at doctor–patient communication grew markedly. Effective communication between providers and patients is very important to the provision of safe and high-quality healthcare. Failures in the communication process can have significant implications for patients’ experience, negatively affecting patient satisfaction, adherence, resource utilisation, and health outcomes [16]. RIAS is based on social exchange theories and linguistic-based techniques of communication analysis, with a coding framework based on the ‘‘three function model” of medical interviewing [15]. The coding is performed directly from the audio recording without transcription, and data are directly entered into RIAS software. The coders assign each word (or complete thought) spoken by the healthcare provider or patient to mutually exclusive and exhaustive categories. These categories can then be combined to reflect four larger functional groupings, namely data gathering, patient education and counselling, building a relationship, and activating and partnering. For example, in the context of STOP, if coders heard open ended questions in a consultation recording, they would categorise this to ‘task focused communication’, where the adviser is ‘gathering data’ to understand the smoker’s problems [16]. Overall, RIAS offers a practical, functional, flexible, and methodologically rigorous approach for reliably analysing patient and provider interactions in a range of contexts [15].

### Pharmacy staff and service user interviews

To assess acceptability of the STOP intervention, we will conduct interviews with a purposive sample of pharmacy staff from participating pharmacies in the intervention arm only and service users who provided their consent during the STOP trial to be subsequently approached at 6 months about their smoking status [1]. We will put emphasis on capturing regional variation so we will purposefully try to get interview participants from all three recruitment sites in case there are differences that impact on service delivery or service user experience [17]. The STOP Trial recruitment targets are 165 staff and 1200 service users [1]. From this potential participant pool, we feel interviewing samples of approximately 10 staff and 25 service users will be sufficient to reach data saturation, considering the homogeneity of the population in each group [18]. Consenting staff will be approached 5 months after training completion to ask if they are willing to participate in a recorded interview. Face-to-face and telephone interviews will be conducted depending on respondents’ location. Service users will be approached during their 6 month follow up, where long term smoking abstinence is assessed. After completing their 6 month follow up questionnaire, the STOP researcher will ask the service user if they would be willing to participate in a recorded interview to give more detail about their experience of using their local pharmacy’s NHS stop smoking service. These interviews will also be conducted by telephone or face to face, whatever is more convenient for the service user. All the interviews will be conducted by members of the STOP team who were not involved in delivery of the STOP intervention training. The interview questions will be guided by semi-structured topic guides (see table 3).

**Table 3:** Summary of topic guides for pharmacy staff and service user interviews

|  |
| --- |
| **Topic guide for interviews with pharmacy staff** |
| ▪ Barriers to training intervention - non-attendance or dropout<br>▪ Facilitators to training intervention attendance<br>▪ Acceptability of training (probe length, structure, content, timing etc.)<br>▪ Feasibility of delivering intervention in practice (probe things that worked/didn’t work)<br>▪ Barriers to implementing the intervention in practice<br>▪ Pharmacy staff view of client perception of intervention received |
| **Topic guide for interviews with smoker service users** |
| ▪ Overall view of programme<br>▪ How they were approached to take part in programme / how did they feel about it<br>▪ Reasons for completing/ dropping out of the programme<br>▪ How were the sessions delivered – what was covered<br>▪ What bits they found most/least useful<br>▪ Were there things that could have been improved in delivery of programme<br>▪ Views on pharmacy environment |

### Booster visit Field notes

After a pharmacy receives STOP intervention training and has been visited by a simulated client (approximately 3 months), the STOP training facilitator will visit the pharmacy to conduct a booster visit. During this visit, the facilitator will meet with the lead stop smoking adviser and/or STOP local champion for the pharmacy (self nominated during the STOP Training) to discuss the pharmacy’s progress implementing the STOP intervention. This will include gauging presence and usefulness of the STOP promotional materials. The facilitator will also give the pharmacy feedback on their simulated client results. Field notes of interactions from these visits will be captured using a STOP Booster Visit Checklist (Figure 3). These field notes will be later analysed to give insights on the extent to which the target population (pharmacy staff) are participating in the intervention.

**Figure 3:**
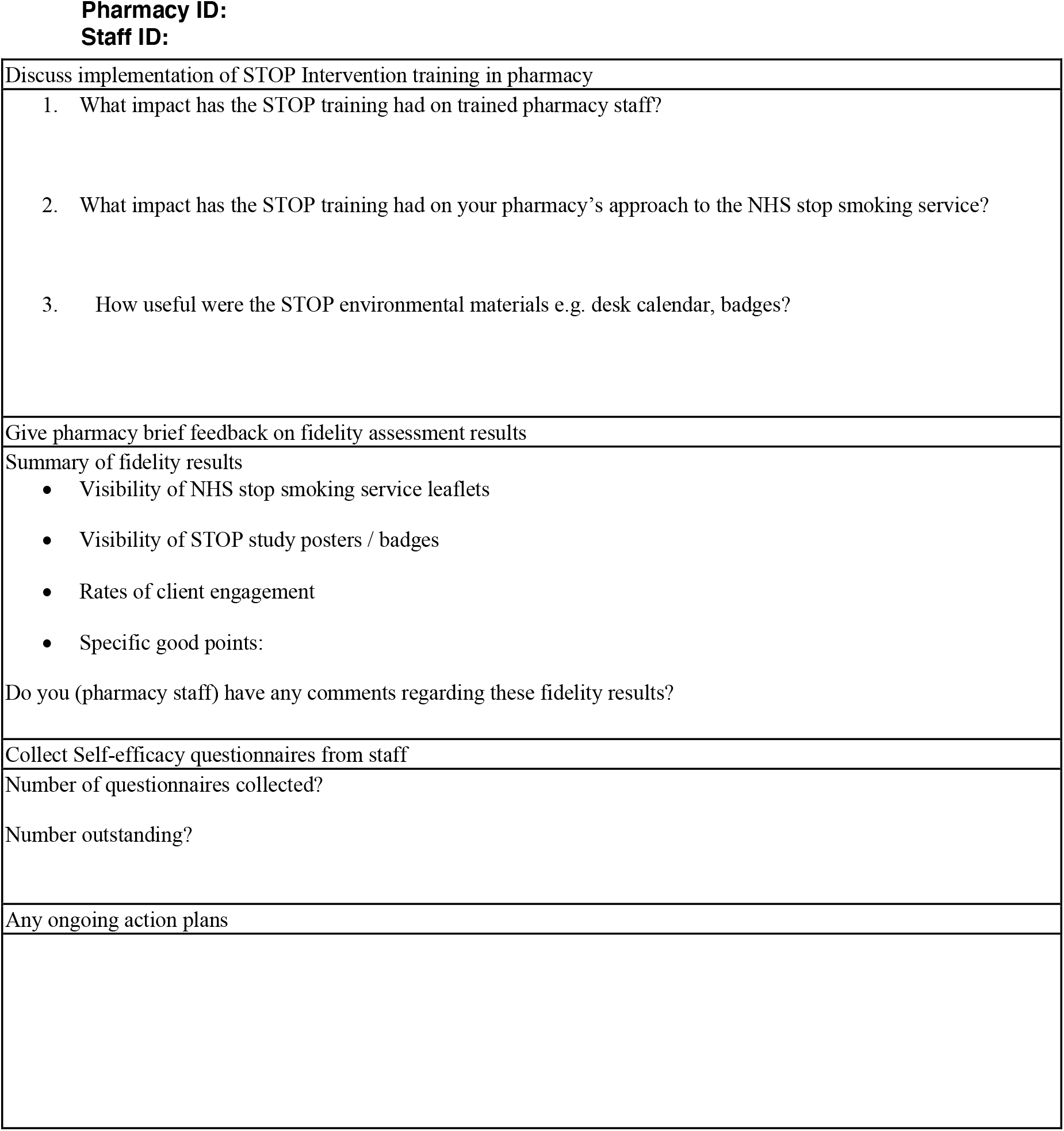
STOP booster visit checklist

### Qualitative Analysis

Interviews and adviser-smoker consultations will be digitally recorded, subject to permission of each participant, and transcribed verbatim by a Queen Mary approved transcription service in line with data management standard operating procedures. The recordings will be stored in a secure virtual environment maintained by our host Centre and will be accessed by authorised study researchers for analysis. Thematic analysis facilitated by the software package NVivo [19] will be used for analysis of the qualitative data. Researcher bias will be minimised through regular crosschecking of data and findings by the members of research team. Anonymised quotations will be used where possible as exemplars of key points in the writing up of these data. Themes emerging from qualitative data will also be discussed and refined by the Process Evaluation Team. To ensure reliability and validity, each team member will code a sample (1-2 transcripts) that have already been coded by another member of the team for subsequent reviewing and reflection as a group on emerging themes.

### Quantitative Analysis

Quantitative data will be analysed using the statistical package PASW Statistics [20]. Descriptive statistics will be generated and comparisons made between intervention and control pharmacies from the mystery shopper data. Regional comparisons will also be examined.

For the training audio-recording data, the first author will summarise total ratings of the pre-classified BCTs. Observed adherence will be expressed as the number of BCTs applied by the facilitator, divided by the number of BCTs specified in the training manual.

The data analysis for the process evaluation data will be an ongoing process and will occur in parallel but independently of the main study, before the two data sets are combined [5]. To ensure good quality data, the process evaluation team will share and review extracted data results at regular intervals.

### Ethics and dissemination

The STOP trial and its process evaluation has ethical approval from the UK National Research Ethics Service (NHS REC reference 17/SC/0067) given on 3 April 2017. Findings will be disseminated via peer reviewed journals and conferences. Considering the limited research and training culture observed in the community pharmacy setting, we also plan to deliver workshops or seminars on methodology of the process evaluation to relevant stakeholders and service commissioning. Brief reports will be shared with participants, mystery shoppers and funder.

## Discussion

This paper describes the design and methodology for the planned process evaluation of the STOP trial using a mixed methods approach. The process evaluation protocol and results will contribute to the continuously developing literature around the conduct of process evaluation for complex health interventions and the value they add to health services research and delivery. The process evaluation protocol follows recommendations intended to help standardise the design and reporting of process evaluation. This will hopefully enable future trials to conduct similar evaluations of their work, resulting in synthesis of stronger evidence base regarding effective interventions.

### Trial status

The current approved STOP Trial protocol is version 4.0 dated 21.03.17. Recruitment to the trial began on 3^rd^ April 2017. Data collection for the process evaluation is still ongoing; to be completed in April 2020.

## Data Availability

The datasets used and/or analysed during the current study are available following completion of the project from the study guarantor, Professor Robert Walton, on reasonable request.

## List of abbreviations

STOP: Smoking Treatment Optimisation in Pharmacies
NHS: National Health Service
BCTs: Behaviour Change Techniques
CPD: Continuing Professional Development
RPS: Royal Pharmaceutical Society
RIAS: Roter Interaction Analysis System
REC: Research Ethics Committee

## Declarations

### Ethics approval and consent to participate

The STOP trial and its process evaluation received ethical approval from South Central - Hampshire A NHS Research Ethics Committee (NHS REC reference 17/SC/0067) on 3^rd^ April 2017

### Consent for publication

“Not applicable”

### Competing interests

“The authors declare that they have no competing interests”

### Funding

This study is funded by the National Institute for Health Research (NIHR) Programme Grants for Applied Research (RP-PG-0609-10181) of which RW is the chief investigator and ST is a co-investigator. The views expressed are those of the author(s) and not necessarily those of the NIHR or the Department of Health and Social Care.

### Authors’ contributions

SJ led on the process evaluation protocol design and acquisition of data with substantial support from CH, SLJ and WYJ. SJ drafted the manuscript and RW, ST and VM substantially contributed to revised versions. All authors have reviewed and commented on the final version.

## Acknowledgements

The authors want to acknowledge all the actors in London, Coventry and Cwm Taf who provided invaluable data from their pharmacy mystery shopper visits. They also thank all the pharmacy staff and service user participants who were willing to be interviewed and audio-recorded.

## Notes

### Competing Interest Statement

The authors have declared no competing interest.

### Clinical Trial

 ISRCTN16351033

